# A Phase I Study of ASTX727 plus Talazoparib in Patients with Triple Negative or Hormone Resistant/HER2-negative Metastatic Breast Cancer

**DOI:** 10.1101/2025.09.09.25335318

**Authors:** Kathy D. Miller, Alexandra Thomas, Sandra Althouse, Yong Zang, Erin Conder, Ryan Burgos, Bryan P. Schneider, Tarah Ballinger, Emily Douglas, Katherine Ansley, H. Josh Jang, Woonbok Chung, Jean-Pierre Issa, Kenneth P. Nephew, Feyruz V. Rassool

## Abstract

**Background:** Poly (ADP-ribose) polymerase inhibitors (PARPi) are effective in patients with germline BRCA 1/2 and PALB2 mutations but have been largely ineffective as monotherapy in others. PARP interacts with, and is recruited to, DNA damage sites along with epigenetic factors, such as DNA methyltransferase 1 (DNMT1). In addition to increasing PARP-trapping, inhibitors of DNMT modulate ROS-cAMP/PKA signaling and induce a pathogen mimicry, inflammasome signaling response and a ‘BRCAness phenotype’ that further sensitizes cells to PARPi. In preclinical *in vitro* and *in vivo* studies, combined DNMTi + PARPi therapy was effective in both triple negative (TNBC) and hormone resistant (HRBC) models with intact BRCA.

**Methods:** We conducted a phase I study combining the oral DNMTi ASTX727 with the PARPi talazoparib in patients (pts) with previously treated TNBC or HRBC; pts with deleterious mutations of BRCA were excluded. Pts with TNBC had received at least one prior chemotherapy and pts with HRBC had received prior endocrine therapy with a cyclin-dependent kinase inhibitor for metastatic disease. An ECOG PS 0-1 and adequate organ function was required. A classical 3+3 design guided dose escalation/de-escalation with dose-limiting toxicity (DLT) defined as Grade 4 neutropenia or thrombocytopenia lasting ≥7 days, or clinically significant grade ≥3 non-hematologic toxicity in cycle 1; 28 days constituted each cycle. Serial peripheral blood mononuclear cells (PBMCs) were analyzed for changes in methylation using the Infinium Methylation EPIC BeadChip and LINE1 sequencing.

**Results:** 34 evaluable pts were enrolled and treated in 8 dose cohorts. Median age was 59 years, 12% identified as Black. Myelosuppression was common with grade ≥3 neutropenia in 42% and grade 3 anemia and thrombocytopenia in 13%. DLT was limited to neutropenia. Efficacy was assessed in 29 pts. There were no objective responses, 6 pts had stable disease persisting for > 4 months in 3 pts. LINE1 demethylation ranged from ∼2-10% and immune-specific CpGs (methylation in immune cells) changed 1-5% by differential methylation locus analysis at Day 15. Methylation changes were not dose dependent.

**Conclusions:** ASTX727 plus talazoparib produces significant myelosuppression but is otherwise well tolerated. Low dose ASTX727 (10 mg decitabine: 100 mg cedazuridine) on Days 1,3,5 with talazoparib 0.5 mg daily on Days 1-21 of each 28-day cycle is recommended for phase II trials. Methylation changes in PBMCs were detected and some heavily pre-treated pts had prolonged stable disease despite the attenuated doses.

**Statement of Translational Relevance:** Inhibitors of poly (ADP-ribose) polymerase (PARPi) have significant clinical benefit in patients with advanced breast cancer who harbor deleterious mutations of BRCA1, BRCA2, or PALB2 but have had limited benefit in those without mutations. Similarly, epigenetic therapies such as the DNA methyltransferase inhibitors (DNMTi) slow disease progression in some hematologic malignancies and myelodysplastic syndromes but have not found a role in solid tumors. Despite limited clinical activity as monotherapy, the combination of PARP and DNMTi significantly inhibited tumor growth in several preclinical models with intact BRCA. In the first clinical trial combining talazoparib and ASTX727, myelosuppression limited drug exposure but the combination was otherwise well tolerated. Strategies to reduce myelosuppression with this combination should be explored. Despite the attenuated doses, methylation changes in peripheral blood mononuclear cells (PMBCs) and clinical benefit were observed; neither were clearly dose dependent.

## Introduction

Triple-negative breast cancer (TNBC) accounts for about 15% of breast cancers. Though immune checkpoint inhibition has a role in some patients^1–5^, chemotherapy remains the mainstay of treatment. Patients with hormone sensitive disease may benefit from an increasing array of antiestrogen regimens^6, 7^, but chemotherapy is recommended when hormone resistance invariably develops. Though multiple chemotherapy agents are available, responses are often short-lived and treatment-related morbidity is substantial. PARP inhibitors (PARPi) have significant activity with minimal toxicity in patients with metastatic breast cancer associated with a deleterious BRCA mutation^8–11^. Patients with deleterious BRCA1 mutations frequently develop TNBC, while those with mutant BRCA2 preferentially develop estrogen sensitive disease. Consequently, there was early hope that PARPi would be useful in sporadic TNBC and hormone resistant disease (HRBC). Unfortunately, clinical reality quickly dashed those hopes, as activity in patients with non-mutated BRCA has been minimal^12, 13^.

Recent investigation of the mechanism underlying the cytotoxic effects of PARPi have shown that the most potent agents trap PARP at DNA damage sites^14–17^, leading to persistent DNA damage and cell death. We have shown that PARP interacts with, and is recruited to, DNA damage sites along with epigenetic factors, such as DNA methyltransferase 1 (DNMT1)^18^. Clinically available PARPis also trap DNMTs into DNA, amplifying the effect. In addition to enzyme trapping, our preclinical data demonstrates that accumulation of reactive oxygen species (ROS) and modulation of the ROS-cAMP/PKA signaling axis by decitabine or guadecitabine significantly enhances the response to talazoparib in TNBC and HRBC cells, resulting in increased sensitivity to talazoparib *irrespective* of BRCA status^16, 18^. More recently, we have shown that the combination of DNMTi and PARPi induces ZNFX1, regulating mitochondrial ROS and leading to mitochondrial damage, DNA leak into the cytosol, and activation of STING similar to the response to viral pathogens^19^. This ‘pathogen mimicry response’ activates inflammasome signaling leading to a ‘BRCAness phenotype’ in models with intact BRCA^20^. We hypothesized that combining DNMTi + PARPi would increase PARP trapping at DNA damage sites, induce ROS, and a ‘BRCAness phenotype’, leading to cytotoxicity in TNBCs and HRBCs with intact BRCA. A phase I trial of decitabine plus talazoparib in patients with refractory acute myelogenous leukemia (AML) was ongoing (NCT02878785)^21^, but there was no experience with DNMTi + PARPi combination regimens in patients with solid tumors.

Our preclinical studies were conducted with the parent DNMTi decitabine and its analogue guadecitabine. Decitabine requires intravenous administration due to rapid clearance by cytidine deaminase (CDA) in the gut and liver^22^. Guadecitabine (2’-deoxy-5-azacytidylyl-(3’→5’)-2’- deoxyguanosine sodium salt) is metabolized to decitabine but is resistant to modification by cytidine deaminase resulting in a longer half-life^23^. However, guadecitabine requires subcutaneous administration and its clinical future was uncertain. Consequently, we chose ASTX727, a novel fixed dose combination of decitabine with cedazuridine, a cytidine deaminase inhibitor with excellent oral bioavailability. A phase I dose finding study found that a fixed dose oral combination of 35 mg of decitabine and 100 mg of cedazuridine produced similar decitabine exposure as decitabine administered intravenously at 20 mg/m2 as a 1-hour infusion. In a confirmatory phase II study, ASTX727 (35:100) successfully emulated the AUC exposures and LINE-1 demethylation of 20 mg/m2 IV decitabine in a 5 consecutive day regimen; clinical response and safety data appeared similar to that reported for decitabine 20 mg/m2 IV as well^24, 25^. In addition, the Day 1, 3, 5 schedule maintained DNMTi. A low dose ASTX727 formulation containing 10 mg decitabine with 100 mg cedazuridine became available during the conduct of this trial, providing greater flexibility in dosing. Given the potential mechanisms of synergy, we selected talazoparib as the PARPi for this trial to maximize PARP trapping. Use of ASTX772 with talazoparib allowed us to test our underlying hypothesis with an all-oral regimen that would be preferred by patients and would facilitate chronic therapy.

## Patients and Methods

### Patient Eligibility

Patients with histologically or cytologically confirmed TNBC or hormone positive, HER2negative metastatic breast cancer were eligible. Disease phenotype was defined based on ASCO-CAP guidelines except that patients with weak ER and PR staining in <5% of cells were considered TNBC. Patients with TNBC must have had at least one prior chemotherapy regimen for metastatic disease; patients with HRBC were required to have progressed on endocrine therapy with a CDKi in the metastatic setting. Patients had to have an ECOG performance status of 0-1 with adequate hepatic (total bilirubin < upper limit of normal (ULN) unless documented Gilbert’s disease; AST and ALT < 3.0 ULN) and hematologic (absolute neutrophils > 1500/mm^3^; platelets > 100,000/ mm^3^; hemoglobin ≥ 9.0 g/dL) function. Measurable or evaluable disease was allowed. Patients with active CNS disease were excluded, but stable CNS involvement (> 4 weeks from definitive CNS treatment with stable or decreasing corticosteroid dose) was allowed. Patients with known deleterious mutations of BRCA1, BRCA2, or PALB2 were excluded. The Indiana University and Wake Forest University Institutional Review Boards reviewed and approved the protocol; all subsequent protocol amendments,all serious adverse events, and annual progress reports. All patients provided individual written informed consent prior to screening and study entry.

### Treatment Plan

Patients were enrolled and treated with oral ASTX727 and talazoparib in eight successive dose cohorts (**Table 1**). Both agents were administered orally once daily; days of administration varied based on the cohort. Patients were asked to fast 2 hours before and 2 hours after ASTX727 administration; clear liquids such as water, black coffee, or tea were allowed during the 4-hour fasting period. Talazoparib was administered without regard to fasting. Patients continued treatment until disease progression or unacceptable toxicity. Prophylactic antiemetics were not recommended but could be added at the investigator’s discretion. Prophylactic use of white or red blood cell growth factors was not allowed but could be added to manage toxicity in accordance with ASCO guidelines, if needed. Concurrent denosumab or bisphosphonates were allowed in patients with bone involvement.

**Table 1.** Dose cohorts.

|  |  | ASTX-727 |  | Talazoparib |  |  |
| --- | --- | --- | --- | --- | --- | --- |
| Dose level | Pts | Decitabine:cedazuridine | Days | Dose | Days | DLTs* |
| 1 | 3 | 35 mg:100 mg | 1-5 | 0.5 mg | 1-28 | 2 |
| 2 | 3 | 35 mg:100 mg | 1-3 | 0.5 mg | 1-28 | 2 |
| 3 | 3 | 35 mg:100 mg | 1,3,5<br>and<br>15,17,19 | 0.5 g | 1-7 and<br>15-21 | 3 |
| 4 | 2 | 35 mg:100 mg | 1,3,5<br>and<br>15,17,19 | 0.25 mg | 1-7 and<br>15-21 | 1 |
| 5 | 6 | 10 mg:100 mg | 1,3,5 | 0.5 mg | 6-21 | 0 |
| 6 | 7 | 10 mg:100 mg | 1-5 | 0.5 mg | 6-21 | 1 |
| 7 | 3 | 10 mg:100 mg | 1,3,5 | 0.25 mg | 1-21 | 0 |
| 8 | 7 | 10 mg:100 mg | 1,3,5 | 0.5 mg | 1-21 | 0 |

\*All DLTs were myelosuppression, primarily neutropenia.

### Safety and Efficacy Assessments

Toxicity was assessed based on NCI Common Terminology Criteria for Adverse Events (CTCAE) version 5.0. Patients were evaluated for dose-limiting toxicity (DLT) defined as Grade 4 neutropenia or thrombocytopenia lasting ≥7 days, or clinically significant grade ≥3 non-hematologic toxicity in cycle 1; 28 days constituted each cycle. Based on the mechanism of action, single agent toxicities, and experience combining DNMTi’s with cytotoxic chemotherapy, myelosuppression was expected. Consequently, a complete blood count was obtained weekly during the first 2 cycles, on Days 1 and 15 of cycle 3, and Day 1 of each subsequent cycle. Doses were held or reduced for excess myelosuppression. Patients were evaluated clinically, and serum chemistries were obtained On Days 1 and 15 of the first 3 cycles, and Day 1 of each subsequent cycle. Disease status was assessed according to the Response Evaluation Criteria for Solid Tumors version 1.1^26^ every two cycles for the first 24 weeks and every 3 cycles thereafter.

### Changes in peripheral blood mononuclear cell (PBMC) methylation

PBMCs were collected for assessment of global DNA methylation of LINE-1 and other selected genes at baseline, prior to treatment on Cycle 1 Day 8, Cycle 2 Day 1, Cycle 2 Day 8, and then Day 1 of Cycles 3, 5, and 7. Genome-wide methylation profiling was conducted using the Infinium HumanMethylationEPIC v2.0 BeadChip platform (Illumina, California, USA). This platform detects methylation levels at over 935,000 CpG sites. Assay process included DNA hybridization to bead arrays, enzymatic extension, and fluorescent staining, enabling precise measurement of methylation at individual sites. We generated DNA methylome data from PBMCs collected at baseline (C1D1), Cycle 1 Day 15 (C1D15), Cycle 2 Day 1 (C2D1), and Cycle 2 Day 15 (C2D15). The raw methylation data were processed with the SeSAMe (Signal Extraction and Summarization of Array Methylation Experiments) R package. This software normalized signal intensities, corrected for dye bias, and filtered low-quality probes, ensuring the reliability of the resulting β-value matrices. These β-values, which range from 0 (unmethylated) to 1 (fully methylated). Furthermore, deconvolution analysis of the methylation data was conducted by HEpiDISH algorithm.

### Bisulfite repetitive element PCR

In addition to methylation profiling, high resolution analysis was performed to assess global DNA methylation changes using our previously published method^27^. Briefly, genomic DNA was isolated from PBMCs (n=62 samples; corresponding to 21 patients studied on day 1, day 15 and C2D1), bisulfite treated, and PCR amplified in non-stringent conditions using PCR primers designed from a consensus LINE repetitive element sequence that allows the amplification of a pool of several thousand repeats. To assess the decrease in global DNA methylation, the sequence difference in this pool of amplified repeats was quantitated using pyrosequencing. In addition, to extend demethylation studies from LINE1 to single loci, bisulfite repetitive element PCR was used to analyze methylation of INS6 (also known as incretin peptide 6), which we have previously shown to have a normally highly methylated promoter CpG island^27^.

### Statistical and Bioinformatic Analyses

The primary objective of this phase I trial was to evaluate the safety and tolerability of ASTX727 in combination with talazoparib in patients with metastatic TNBC or HRBC. Three patients were enrolled into each successive cohort. If none of the 3 patients in a cohort experienced DLT, accrual to the next cohort commenced. If 2 or more patients experienced DLT, the previous dose level was considered maximum tolerated dose (MTD). If DLT was observed in 1 of 3 patients, 3 additional patients were to be added to that dose level. If no additional DLTs were reported, accrual to the next cohort commenced. If ≥ 2 of the 6 patients experienced DLT, the previous dose level was considered the MTD. Patients who stopped therapy before completing the DLT evaluation period for reasons other than toxicity were replaced. The secondary endpoint was to evaluate efficacy; circulating biomarkers of response and biologic activity were exploratory endpoints. Demographic and other characteristics were summarized as median (range) for continuous variables and number and percentage for categorical variables. Overall Response Rate (ORR) and Clinical Benefit Response (CBR; CR/PR/SD at 18 weeks for TNBC, 24 weeks for HRBC) were determined and the percentage and 95% confidence intervals were calculated, if possible. The Kaplan-Meier method was used to analyze progression free survival (PFS) and overall survival (OS). Median with 95% confidence intervals were calculated along with the 6-month probability. All analyses were performed using SAS Version 9.4 (Cary, NC).

## Results

Thirty-four patients were enrolled and evaluable for toxicity. Twenty-nine patients were evaluable for response; one did not have measurable disease at baseline and four stopped treatment without post-baseline disease assessment. Median age was 59 years (33-76). Most patients were White (85%); 12% identified as Black. Three-quarter (73.5%) of patients had an ECOG performance status of 1 at study entry.

As expected, myelosuppression was common with grade ≥3 neutropenia in 42% and grade ≥ 3 anemia and thrombocytopenia in 13% (**Table 2**); three patients (8.8%) stopped therapy due to myelosuppression. DLT was limited to neutropenia (**Table 1**). Most patients (79%) stopped therapy due to disease progression; one patient died due to disease progression within 30 days of her last study therapy. There were no treatment related deaths.

**Table 2.** Treatment-related Adverse Events*.

|  | Grade 2<br>N (%) | Grade 3<br>N (%) | Grade 4<br>N (%) |
| --- | --- | --- | --- |
| Neutropenia | 2 (6) | 3 (9) | 11 (32) |
| Neutropenic fever | 0 | 3 (9) | 1 (3) |
| Thrombocytopenia | 1 (3) | 4 (12) | 1 (3) |
| Anemia | 7 (21) | 5 (15) | 0 |
| Fatigue | 8 (24) | 1 (3) | 0 |
| Mucositis | 2 (6) | 0 | 0 |
| Hyponatremia | 2 (6) | 1 (3) | 0 |
| Increased AST | 2 (6) | 0 | 0 |
| Insomnia | 3 (9) | 0 | 0 |
| Hypokalemia | 1 (3) | 1 (3) | 0 |
| Infection (not neutropenic) | 1 (3) | 1 (3) | 0 |
| Hypocalcemia | 1 (3) | 1 (3) | 0 |

\*Includes all events with Grade 2 or greater toxicity in at least 2 patients.

There were no objective responses, 6 pts had stable disease persisting for > 4 months in 3 pts with ER+ disease (**Figure 1**). No patients achieved a clinical benefit response as defined *a priori*. Median PFS was 1.7 months (1.6, 1.8); six-month PFS probability was 6.7% (1.2, 19.2). Median overall survival was 7.3 months (4.6, 8.2).

**Figure 1.**
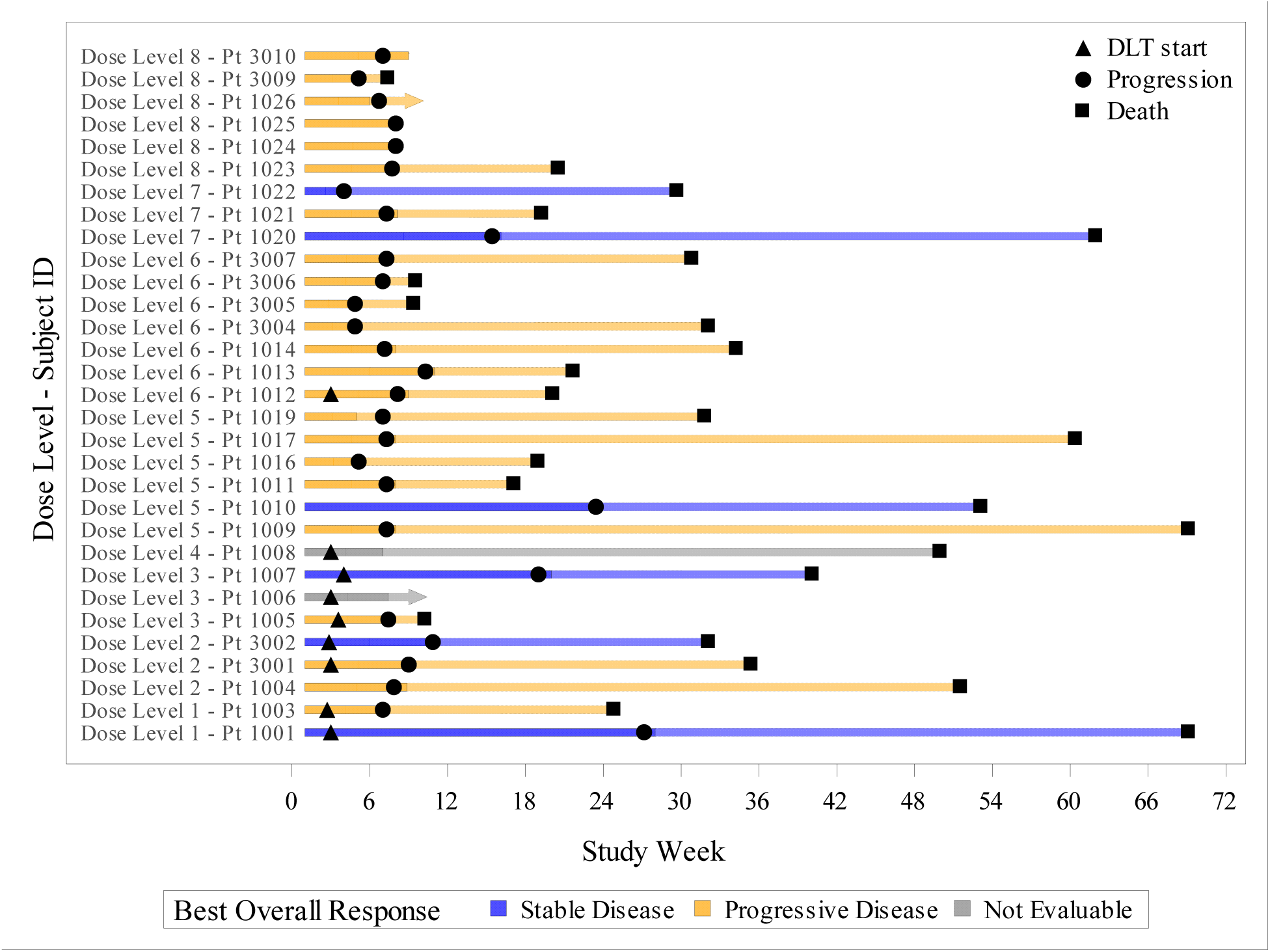
Swimmer plot documenting treatment exposure, DLT, and response. DLTs are indicated by a triangle, disease progression by a circle, and death a square. Patients with a best response of stable disease are shown in yellow and those with progressive disease in purple.

Overall LINE demethylation ranged from 5-10%, based on genome-wide methylation profiling using all CpG probes (**Figure 2A, u**pper), intergenic LINE probes (**Figure 2A, lower**) and the change in median DNA methylation levels in PBMCs 15 days after the start of the treatment cycle (**Figure 2B**). Although methylation changes were not dose dependent, high resolution analysis revealed variable LINE1 demethylation in different doses/cohorts (**Figure 3A, B**). For example, more pronounced demethylation was observed in two cohorts (**Figure 3B**, Cohorts 3 (green line) and 6 (orange line). As expected, there was a trend for the sequential regimens to have more demethylation (8.3%) compared to the concurrent dosing cohorts (5.1%), despite a much lower dose of ASTX727 (44 mg/cycle vs 123 mg/cycle). Although this difference was not significant, the observation provides evidence for an interaction between the two drugs. In the sequential regimen, demethylation seen in Cohort 5 (2.6%, n=2, purple line) compared to Cohort 6 (10.6% demethylation, n=5, orange line), but the number of patients was small.

**Figure 2.**
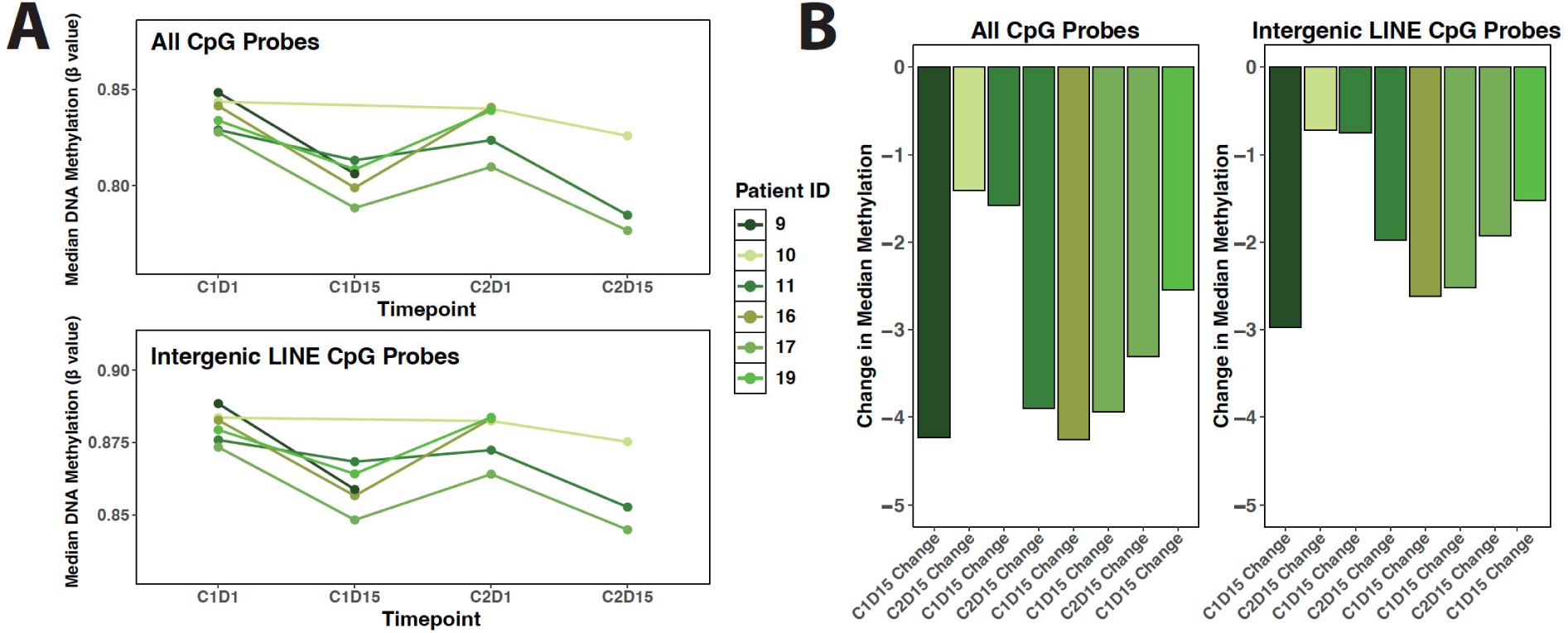
Global loss of DNA methylation in patient PBMCs following treatment. **A**) LINE graphs showing the median DNA methylation levels in patient PBMC samples, assessed using all CpG probes or intergenic LINE CpG probes, across treatment timepoints. **B**) Bar plots depicting the change in median DNA methylation levels in PBMCs 15 days after the start of the treatment cycle.

**Figure 3.**
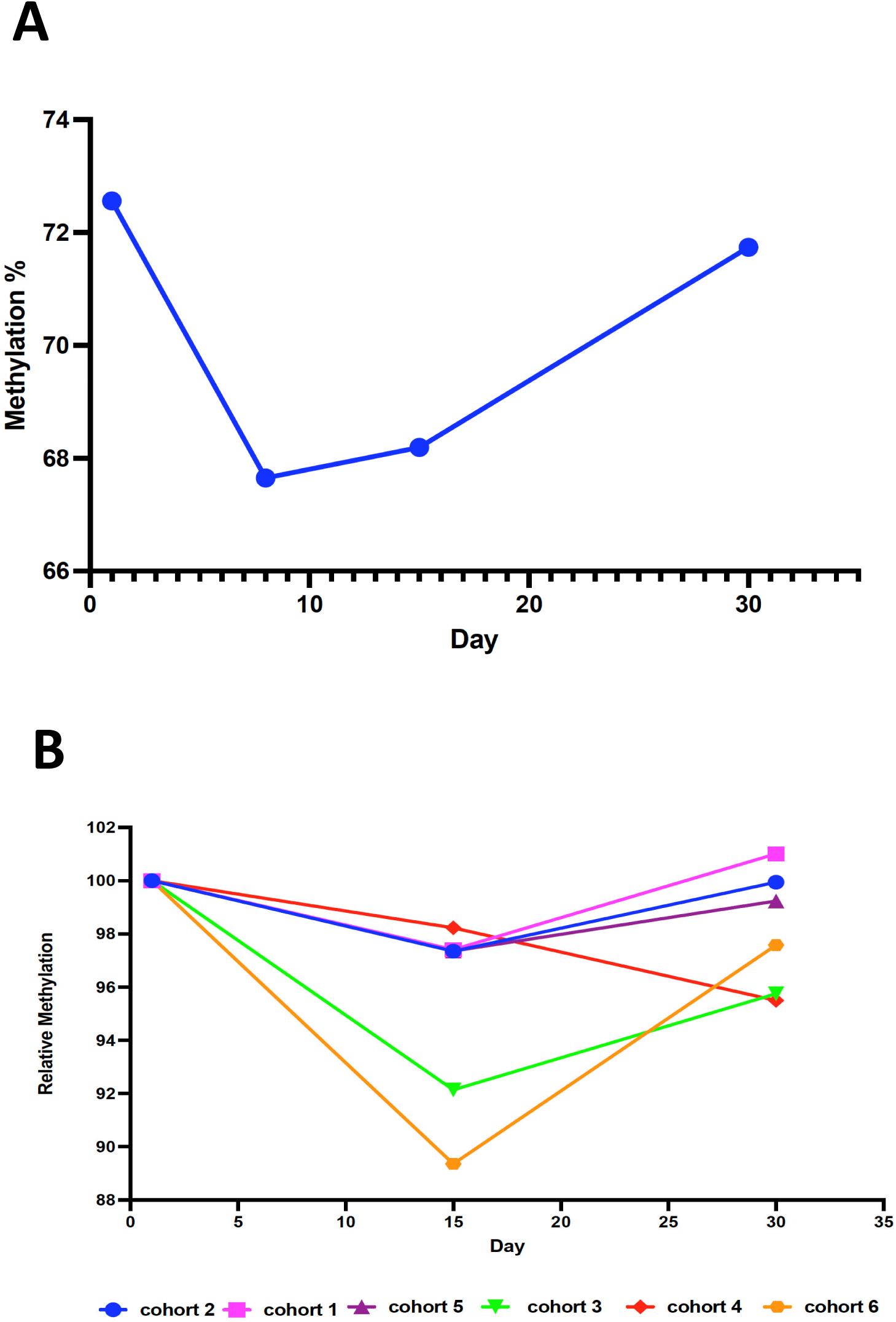
Methylation levels of LINE1 repetitive element. LINE1 was used to to assess global DNA methylation changes in PBMCs isolated from **A)** baseline and prior to treatment on different time points or **B)** collected from the cohorts described in Table 1.

INS6, which has a normally highly methylated promoter CpG island, was used to extend demethylation studies from LINE1 to single loci. All 21 cases were examined. INS6 demethylation varied across cohorts, ranging from <1% (e.g., Cohort 5) to >11% (e.g., Cohort 6) (**Supplemental Figure S1**). INS6 demethylation correlated highly with LINE1 (R2=0.8 for the LINE1 shown in Figure 3). It should be noted that LINE1 is more sensitive to demethylation (demethylates 2-3 times more than INS6) and that no samples showed increased methylation of INS6, confirming assay validity.

Deconvolution analysis of the methylation data demonstrated global and intergenic LINE CpGs changed ∼1-4% by differential methylation locus analysis at Day 15 (**Figure 4 A, B**). Violin plots comparing of β-value ranges for CpG site methylation levels pre- and post-treatment demonstrated consistent bimodal distributions, with peaks near 0 and 1 corresponding to hypo- and hypermethylated regions (**Supplemental Figure S2).** There was a general trend (pre vs. post-treatment) toward loss of monocytes and gain of CD8 and CD4 T cells (**Figure 4**).

**Figure 4.**
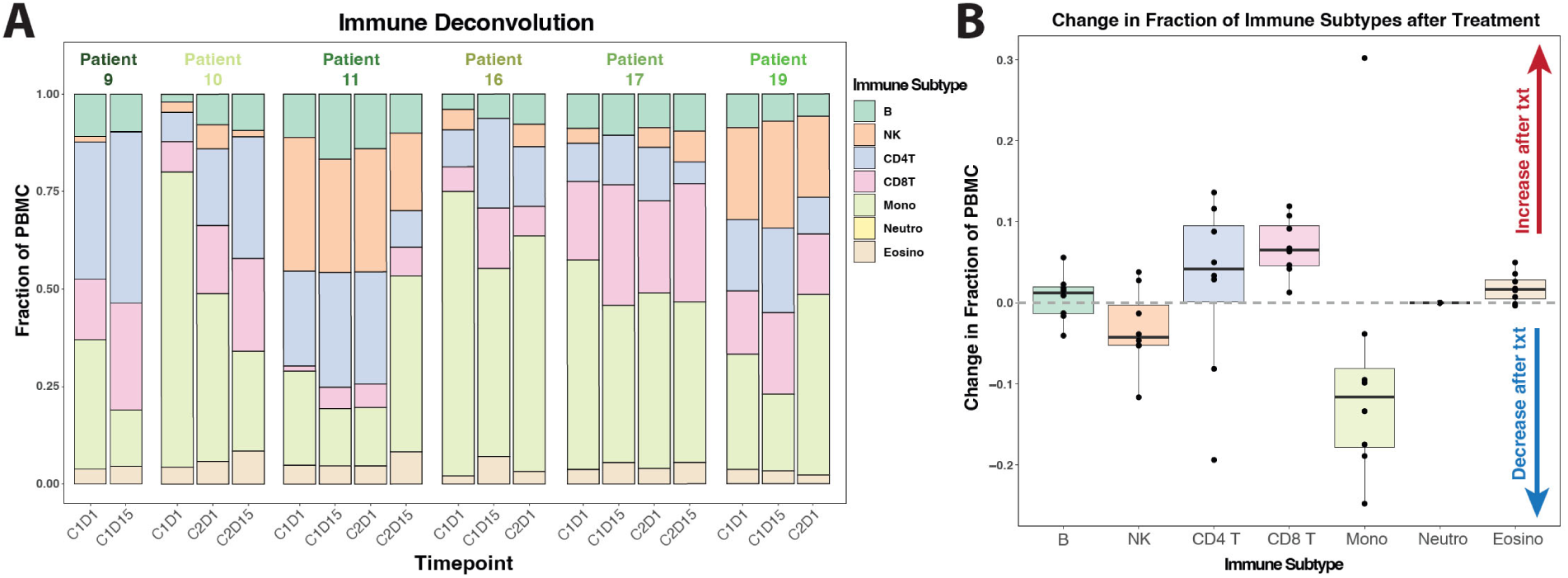
Immune cell dynamics in patient PBMCs following treatment. **A)** Immune cell subtype proportions inferred from PBMC samples using EpiDISH deconvolution analysis. **B)** Box plots showing changes in immune cell fractions 15 days after the start of the treatment cycle.

## Discussion

A previous phase I trial explored the combination of decitabine and talazoparib in patients with AML unfit for or resistant to cytotoxic chemotherapy. That trial enrolled 25 patients over 7 cohorts, ultimately arriving at a recommended phase II dose of decitabine 20 mg/m2 intravenously daily for 5 or 10 days and talazoparib 1 mg orally daily for 28 days. Two patients achieved complete remission with incomplete count recovery; three had hematologic improvement. Pharmacodynamic studies showed the expected DNA demethylation and increased PARP trapping in chromatin. γH2AX foci increased significantly with increasing talazoparib doses combined with 20 mg/m2 decitabine^21^. However, given the nature of AML, hematologic toxicity could not be evaluated in that trial, prompting our first phase I trial of a DNMTi with a PARPi in patients with solid tumors. Myelosuppression was not predicted by the preclinical models and was even more profound than we had anticipated, leading to multiple amendments to explore alternate dosing strategies.

Though we were able to identify a safe and tolerable dose, real questions remained as to whether doses were below the level needed for biologic and clinical activity. We did detect changes in methylation in PBMCs, but the changes were small and not clearly dose dependent. The correlative analysis is limited by the small samples size and focus on PBMCs. To facilitate rapid accrual, we did not collect serial tumor biopsies so could not assess changes in methylation in tumor samples. However, work in other settings have found a poor correlation between methylation changes in PBMCs and tumor samples^28–32^. Despite the dose attenuation, we did see some signs of clinical activity in previously treated patients. PARP1-specific inhibitors may have less myelosuppression than the broad inhibitors currently in clinical use^33–36^. If synergy is maintained, the combination of a PARP1-specific inhibitor with a DNMTi would be worth investigating.

While we focused on models and patients without BRCA mutations, the synergy may also amplify the benefit of PARPi’s in patients with deleterious mutation of BRCA or other genes in the homologous recombination (HR) pathway^37^. The combination of ASTX727 and olaparib is being studied in an ongoing phase I/II trial (NCT06177171) in patients with advanced/metastatic solid tumors and germline or somatic mutations in genes involved in homologous recombination (i.e., BRCA1, BRCA2, PALB2, ATM, or CHEK2). In addition to defining the maximum tolerated dose and evaluating clinical response, exploratory correlative studies will characterize the various homologous recombination pathway mutations and their functional implications and create patient derived xenografts and organoids to study resistance mechanisms.

The synergistic effect could also be maintained by combining PARPi’s with direct STING agonists^38^. Pedretti and colleagues analyzed 35 breast cancer patient-derived xenografts (PDX) and mouse-derived allografts (MDA). The cGAS-STING-IFN pathways was activated in tumors sensitive to PARPi. The combination of PARPi and a novel STING agonist (STINGa) increased immune infiltration and antitumor activity. Of note, additional analyses highlighted the importance of NK cell engagement, further supporting combination with immune therapies^38^.

Other mechanisms to expand the potential benefit of PARPi to patients with intact HR have also been studied^39, 40^. The combination of everolimus and niraparib was not feasible even at reduced doses due to rapid onset and severe hypertension^41^. A phase 1b dose escalation and expansion study of the pan-PI3Ki buparlisib (BKM120) and olaparib found clinical benefit in patients with and without germline BRCA mutations but required attenuation of the buparlisib dose due to toxicity^42^. The alpha-specific Pi3K inhibitor alpelisib has also been successfully combined with olaparib. Of the 28 patients with epithelial ovarian cancer, ten (36%) achieved a partial response and 14 (50%) had stable disease^43^. In 17 patients with previously treated TNBC, ORR was 18% (23% for patients treated at the recommended phase II dose)^44^. The combination of olaparib and the AKT inhibitor capivasertib was well tolerated in a recently reported phase I trial. Pharmacodynamic studies confirmed phosphorylated (p) GSK3beta suppression, increased pERK, and decreased BRCA1 expression. Antitumor activity was observed in patients harboring tumors with germline BRCA1/2 mutations and BRCA1/2 wild-type cancers with or without somatic DDR and/or PI3K-AKT pathway alterations^45^. We recently conducted the first study to combine talazoparib with an mTOR/pan-PI3K inhibitor (gedatolisib) mTOR/PI3 Ki patients with advanced triple negative breast cancer or advanced HER2 negative breast cancer and a germline BRCA1/2 mutation^46^. The combination was safe but did not meet its primary efficacy response threshold. HRD status (measured by genomic instability) was not correlated with response, suggesting functional HRD assessments may need to be examined.

To better understand the effect of hypomethylating agents on immune cells, we examined DNA methylation changes in patient PBMCs. LINE1 PBMC methylation levels decreased after ASTX727 treatment, and global hypomethylation of CpG loci in PBMCs was also observed. However, an unexpected finding of altered components of PBMCs by ASTX727, including monocytes and lymphocytes, was apparent, and the observed gain in T cells could reflect an altered (activated) tumor microenvironment, one more susceptible to immunotherapy.

This is the first report on the global impact of a hypomethylating agent-PARPi combination treatment on PBMC methylomes in breast cancer patients. While the effect of the low-dose ASTX727 with talazoparib recommended for phase II trials on PBMC methylation levels was modest, it is also possible that the hypomethylation effect could be masked by an increased cytotoxic activity of talazoparib on demethylated cells. Further studies combining demethylating agents and PARPi are warranted.

## Data Availability

All data produced in the present study are available upon reasonable request to the authors

---

**Supplementary Figure 1.**
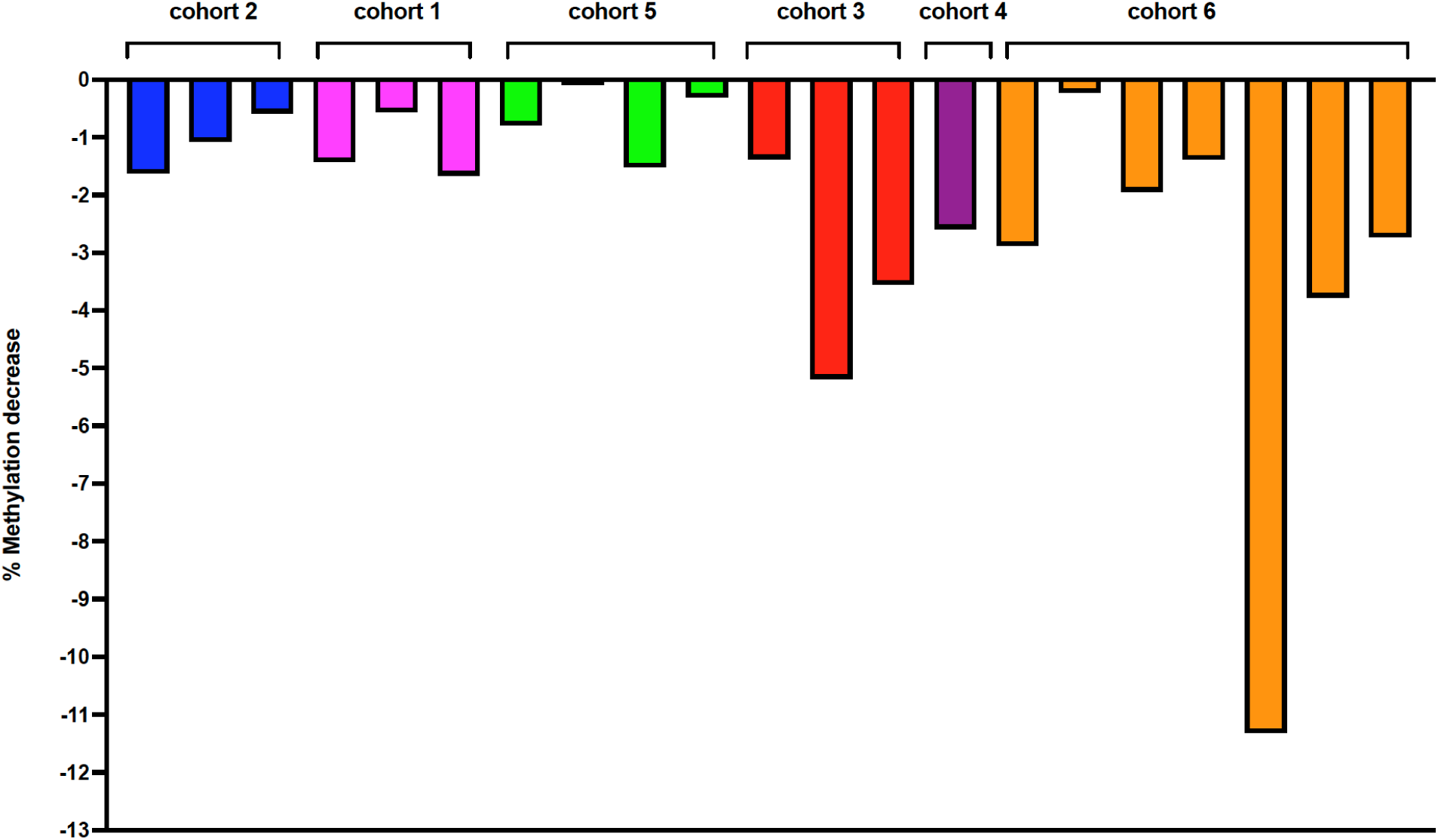
Methylation levels of incretin peptide 6 (INS6) in patient PBMCs. DNA methylation levels were measured using bisulfite repetitive element PCR and primers specific for INS6.

**Supplementary Figure 2.**
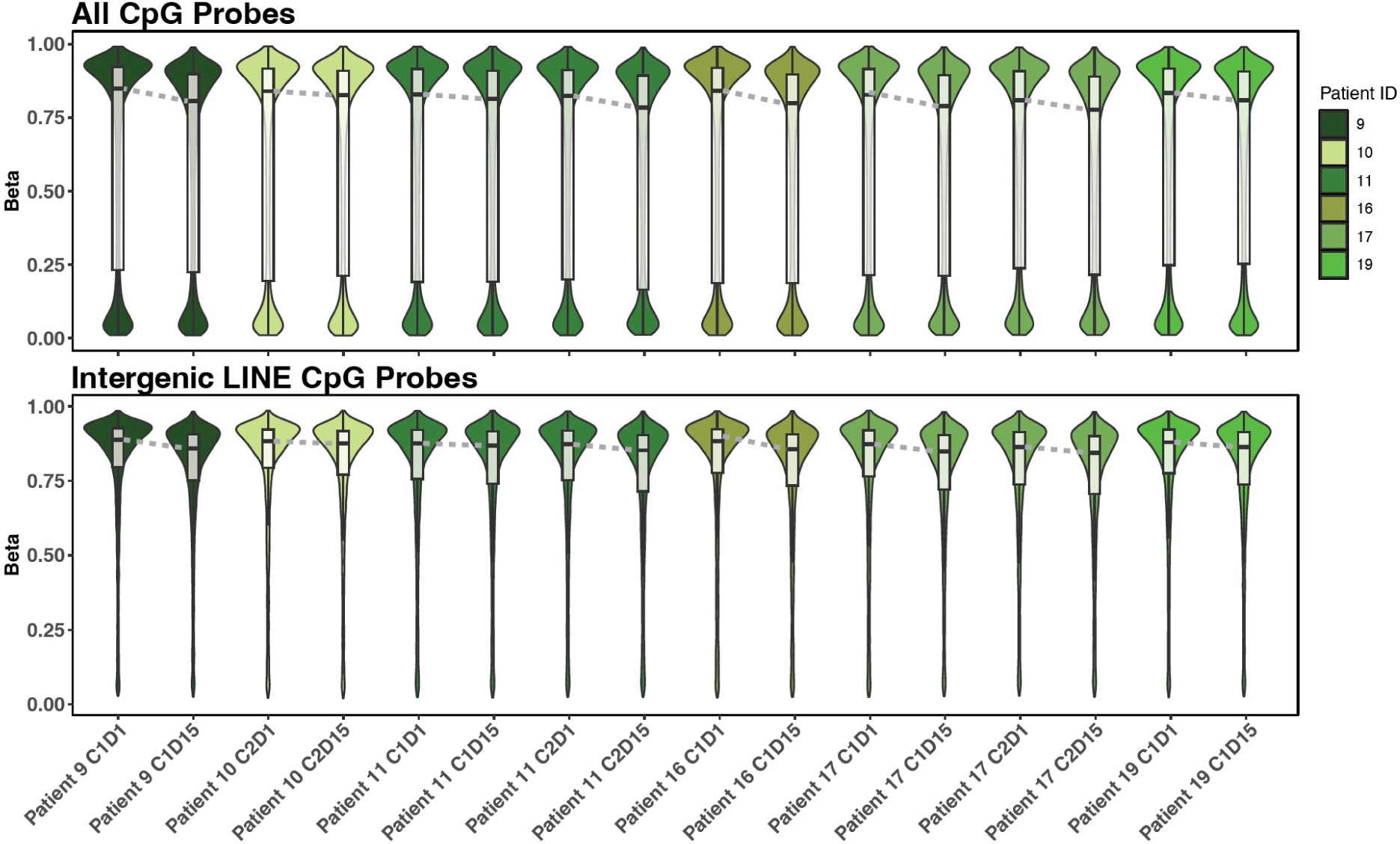
Distribution of CpG methylation levels in patient PBMCs across treatment timepoints. DNA methylation levels measured using all CpG probes or intergenic LINE CpG probes in PBMC samples collected at different timepoints during treatment.

